# COVID-19 testing among children, parental preferences for testing venues and acceptability of school-based testing: a survey of US parents

**DOI:** 10.1101/2021.05.27.21257932

**Authors:** Chloe A. Teasdale, Luisa N. Borrell, Yanhan Shen, Spencer Kimball, Michael L. Rinke, Sasha A. Fleary, Denis Nash

**Author notes:** **Corresponding author**: Chloe A. Teasdale, PhD, Assistant Professor, Department of Epidemiology & Biostatistics, CUNY Graduate School of Public Health & Health Policy, 55 W125th Street, Room 543, New York, NY 10025. **Funding**: This study was funded internally by the Institute for Implementation Science in Population Health (ISPH) of the City University of New York (CUNY) Graduate School of Public Health and Health Policy (SPH). The authors declare no conflicts of interest.

## Abstract

In a national survey of 2,074 US parents conducted in March 2021, 35.9% reported their youngest child had been tested at least once for COVID-19. Parents’ preferred testing venue choice was the pediatrician’s office. Only half of parent surveyed (50.6%) reported that they would allow their child to be tested for COVID-19 at school/daycare if it was required.

---

Approximately 3.9 million children in the United States (US) have been diagnosed with COVID-19 since the start of the epidemic and there have been close to 300 pediatric deaths (as of May 13, 2021).(1) The absolute number of COVID-19 cases in the US continues to decline since the peak in January 2021. However, starting in April 2021, infections in children have been increasing as a proportion of all newly diagnosed cases of COVID-19 and may be contributing to surging infections in parts of the US.(2) According to the American Academy of Pediatrics (AAP), since the start of the epidemic, children have represented 14.0% of all newly diagnosed COVID-19 cases whereas in the first week of May, 2021 they were 24.0% (72,067 new cases identified in children).(1)

While most children infected with COVID-19 experience mild symptoms and have markedly lower mortality compared to adults, COVID-19 infection can lead to multi□inflammatory syndrome in children (MIS□C) and new evidence suggests that some children may experience persistent symptoms after infection.(3, 4) Diagnosing infections in children is critical for providing appropriate care and for helping to contain the spread of COVID-19, including in school and daycare settings.(5) Children however remain a low proportion of all those tested in the US. Data from the first week of May, 2021 show that across 11 states, out of all COVID-19 tests performed, the proportion of tests in children ranged from 6.0% to 19.4%.(1) There have been few studies examining testing coverage among children and routine surveillance generally only captures information on age with few other characteristics reported.(6)

To provide more in-depth information about COVID-19 testing in pediatric populations across the US and to facilitate planning, we conducted an online survey of parents to determine the proportion of children who have ever been tested. We also identified the preferred testing venues and whether parents would allow their child to be tested for COVID-19 at school or daycare.

## METHODS

We conducted an online national survey of US parents and caregivers (‘parents’) using a Qualtrics panel recruited through social media and partner networks. Eligible participants were English and Spanish speaking adults ≥18 years identifying as a primary caregiver of a child ≤12 years. We followed the American Association for Public Opinion Research (AAPOR) guidelines for quota-based sampling.(7) Survey weights were calculated based on national estimates of US parents according to 2019 US Census data calculated using sex, race, ethnicity, education, and region.(8) Data were collected from March 9 through April 2, 2021. The institutional review board of the CUNY Graduate School of Public Health and Health Policy provided ethical approval.

Parents reported information about the youngest child living in the household. Outcomes were: proportion of parents reporting child tested for COVID-19 (“has your child ever been tested for COVID-19”); where parents would take child for testing (“if your child needs testing for COVID-19 in the future, where would you take him/her to be tested”) including multiple response options; and acceptance of school-based testing (“if your child’s school or daycare required COVID-19 testing on a random basis, would you allow your child to be tested at school or daycare”). Parents also reported demographics and household information.

Survey weights were applied to generate national estimates among US parents. We present unweighted counts and weighted percentages for the sample and cumulative incidence estimates for pediatric testing, preferred testing venues and acceptance of school/daycare testing. Rao adjusted Pearson chi-squared tests were used to compare incidence by characteristics. Poisson regression models with robust standard errors were used to estimate adjusted risk ratios (aRR) of pediatric testing adjusted for demographic and household characteristics.

## RESULTS

Among 2,074 US parents surveyed, 35.9% reported their youngest child (median child age: 4.8 years) had ever been tested for COVID-19 (Table 1). In adjusted models, parents significantly less likely to report a child having been tested for COVID were female (aRR 0.69 95%CI 0.60-0.79), Asian (aRR 0.58; 95%CI 0.39-0.87) and from the Midwest (aRR 0.76; 95%CI 0.63-0.91). Children with health insurance (aRR 1.38; 95%CI 1.05-1.81) and those attending in-person school/daycare (aRR 1.67; 95%CI 1.43-1.95) were significantly more likely to have been tested (Table 1). When asked to select venues where they would take their child for testing, 52.7% of all parents selected pediatrician’s office, 27.9% selected drive through testing site, 24.3% selected hospital, 21.8% selected health department testing site and 20.6% selected urgent care (data now shown).

**Table 1.**
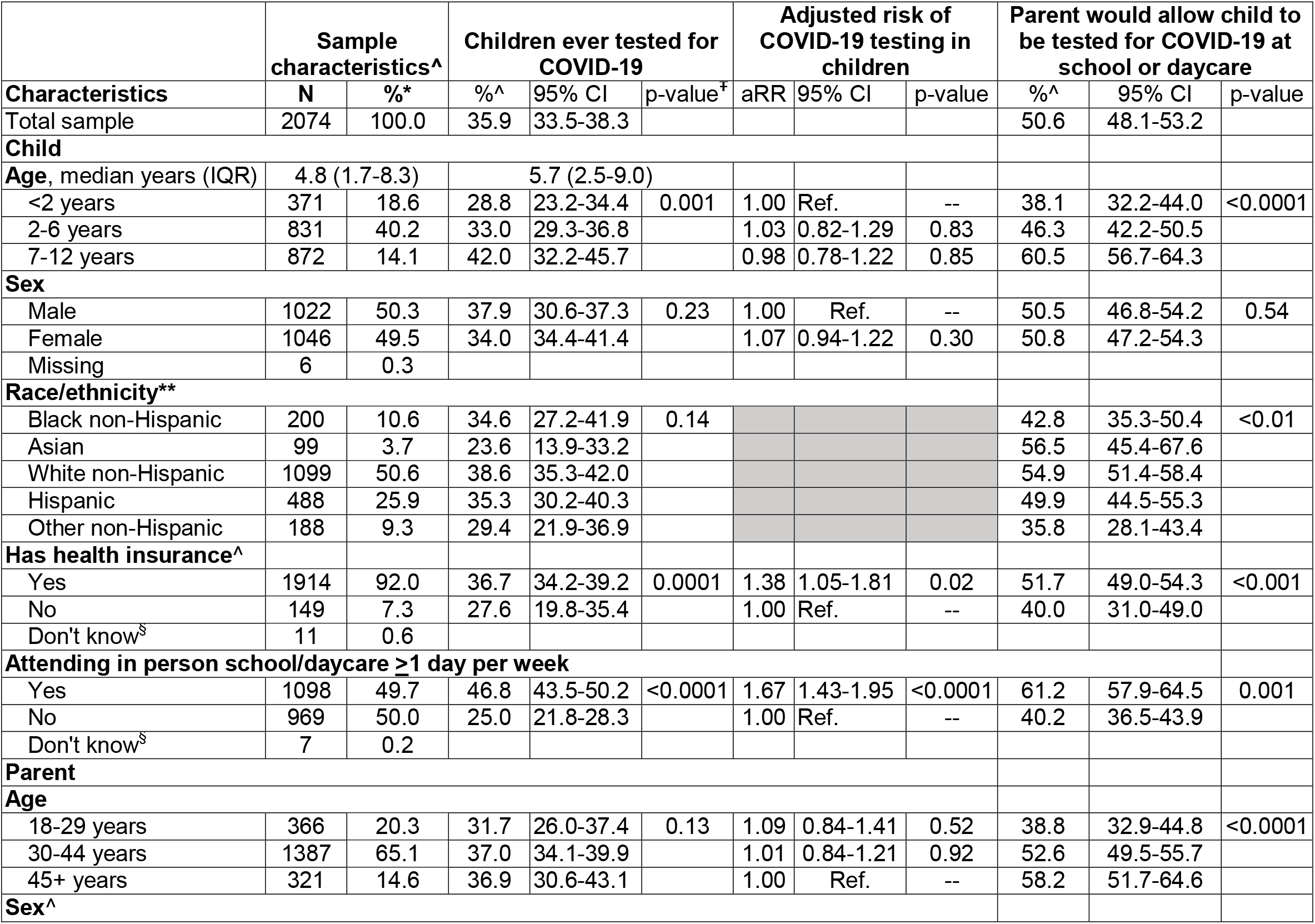
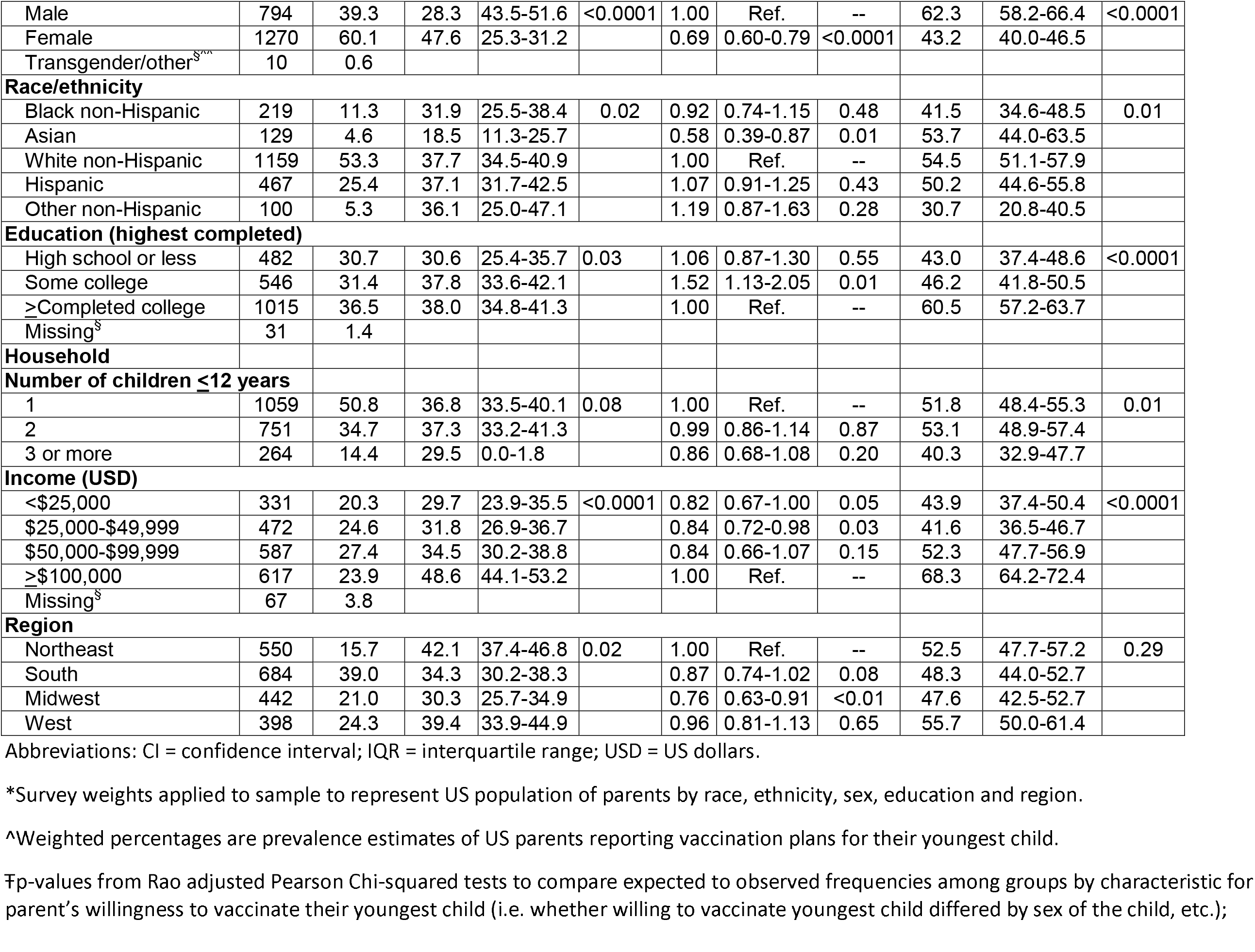
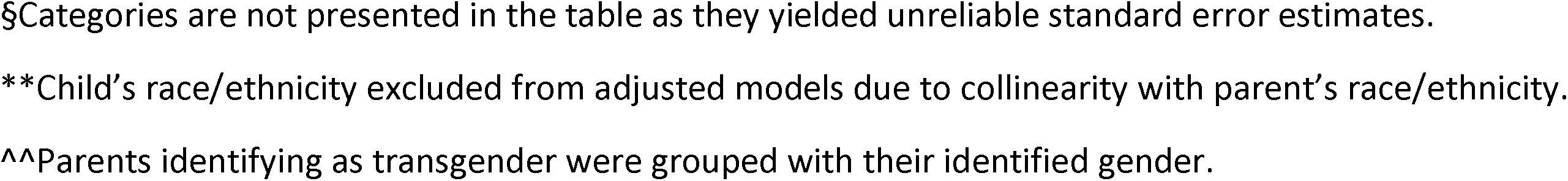
Characteristics of US children ≤12 years and parents, estimated cumulative incidence of testing for COVID-19, adjusted risk ratios for COVID-19 testing (vs. not tested) and prevalence of parental willingness to have children tested at school or daycare, (data collected March 9-April 2, 2021)

|  | Sample characteristics^ |  | Children ever tested for COVID-19 |  |  | Adjusted risk of COVID-19 testing in children |  |  | Parent would allow child to be tested for COVID-19 at school or daycare |  |  |
| --- | --- | --- | --- | --- | --- | --- | --- | --- | --- | --- | --- |
| Characteristics | N | %* | %^ | 95% CI | p-value<sup>†</sup> | aRR | 95% CI | p-value | %^ | 95% CI | p-value |
| Total sample | 2074 | 100.0 | 35.9 | 33.5-38.3 |  |  |  |  | 50.6 | 48.1-53.2 |  |
| **Child** |  |  |  |  |  |  |  |  |  |  |  |
| Age, median years (IQR) | 4.8 (1.7-8.3) |  | 5.7 (2.5-9.0) |  |  |  |  |  |  |  |  |
| <2 years | 371 | 18.6 | 28.8 | 23.2-34.4 | 0.001 | 1.00 | Ref. | -- | 38.1 | 32.2-44.0 | <0.0001 |
| 2-6 years | 831 | 40.2 | 33.0 | 29.3-36.8 |  | 1.03 | 0.82-1.29 | 0.83 | 46.3 | 42.2-50.5 |  |
| 7-12 years | 872 | 14.1 | 42.0 | 32.2-45.7 |  | 0.98 | 0.78-1.22 | 0.85 | 60.5 | 56.7-64.3 |  |
| **Sex** |  |  |  |  |  |  |  |  |  |  |  |
| Male | 1022 | 50.3 | 37.9 | 30.6-37.3 | 0.23 | 1.00 | Ref. | -- | 50.5 | 46.8-54.2 | 0.54 |
| Female | 1046 | 49.5 | 34.0 | 34.4-41.4 |  | 1.07 | 0.94-1.22 | 0.30 | 50.8 | 47.2-54.3 |  |
| Missing | 6 | 0.3 |  |  |  |  |  |  |  |  |  |
| **Race/ethnicity\*\*** |  |  |  |  |  |  |  |  |  |  |  |
| Black non-Hispanic | 200 | 10.6 | 34.6 | 27.2-41.9 | 0.14 |  |  |  | 42.8 | 35.3-50.4 | <0.01 |
| Asian | 99 | 3.7 | 23.6 | 13.9-33.2 |  |  |  |  | 56.5 | 45.4-67.6 |  |
| White non-Hispanic | 1099 | 50.6 | 38.6 | 35.3-42.0 |  |  |  |  | 54.9 | 51.4-58.4 |  |
| Hispanic | 488 | 25.9 | 35.3 | 30.2-40.3 |  |  |  |  | 49.9 | 44.5-55.3 |  |
| Other non-Hispanic | 188 | 9.3 | 29.4 | 21.9-36.9 |  |  |  |  | 35.8 | 28.1-43.4 |  |
| **Has health insurance^** |  |  |  |  |  |  |  |  |  |  |  |
| Yes | 1914 | 92.0 | 36.7 | 34.2-39.2 | 0.0001 | 1.38 | 1.05-1.81 | 0.02 | 51.7 | 49.0-54.3 | <0.001 |
| No | 149 | 7.3 | 27.6 | 19.8-35.4 |  | 1.00 | Ref. | -- | 40.0 | 31.0-49.0 |  |
| Don't know<sup>§</sup> | 11 | 0.6 |  |  |  |  |  |  |  |  |  |
| **Attending in person school/daycare ≥1 day per week** |  |  |  |  |  |  |  |  |  |  |  |
| Yes | 1098 | 49.7 | 46.8 | 43.5-50.2 | <0.0001 | 1.67 | 1.43-1.95 | <0.0001 | 61.2 | 57.9-64.5 | 0.001 |
| No | 969 | 50.0 | 25.0 | 21.8-28.3 |  | 1.00 | Ref. | -- | 40.2 | 36.5-43.9 |  |
| Don't know<sup>§</sup> | 7 | 0.2 |  |  |  |  |  |  |  |  |  |
| **Parent** |  |  |  |  |  |  |  |  |  |  |  |
| **Age** |  |  |  |  |  |  |  |  |  |  |  |
| 18-29 years | 366 | 20.3 | 31.7 | 26.0-37.4 | 0.13 | 1.09 | 0.84-1.41 | 0.52 | 38.8 | 32.9-44.8 | <0.0001 |
| 30-44 years | 1387 | 65.1 | 37.0 | 34.1-39.9 |  | 1.01 | 0.84-1.21 | 0.92 | 52.6 | 49.5-55.7 |  |
| 45+ years | 321 | 14.6 | 36.9 | 30.6-43.1 |  | 1.00 | Ref. | -- | 58.2 | 51.7-64.6 |  |
| **Sex^** |  |  |  |  |  |  |  |  |  |  |  |
| Male | 794 | 39.3 | 28.3 | 43.5-51.6 | <0.0001 | 1.00 | Ref. | -- | 62.3 | 58.2-66.4 | <0.0001 |
| Female | 1270 | 60.1 | 47.6 | 25.3-31.2 |  | 0.69 | 0.60-0.79 | <0.0001 | 43.2 | 40.0-46.5 |  |
| Transgender/other<sup>sxx</sup> | 10 | 0.6 |  |  |  |  |  |  |  |  |  |
| **Race/ethnicity** |  |  |  |  |  |  |  |  |  |  |  |
| Black non-Hispanic | 219 | 11.3 | 31.9 | 25.5-38.4 | 0.02 | 0.92 | 0.74-1.15 | 0.48 | 41.5 | 34.6-48.5 | 0.01 |
| Asian | 129 | 4.6 | 18.5 | 11.3-25.7 |  | 0.58 | 0.39-0.87 | 0.01 | 53.7 | 44.0-63.5 |  |
| White non-Hispanic | 1159 | 53.3 | 37.7 | 34.5-40.9 |  | 1.00 | Ref. | -- | 54.5 | 51.1-57.9 |  |
| Hispanic | 467 | 25.4 | 37.1 | 31.7-42.5 |  | 1.07 | 0.91-1.25 | 0.43 | 50.2 | 44.6-55.8 |  |
| Other non-Hispanic | 100 | 5.3 | 36.1 | 25.0-47.1 |  | 1.19 | 0.87-1.63 | 0.28 | 30.7 | 20.8-40.5 |  |
| **Education (highest completed)** |  |  |  |  |  |  |  |  |  |  |  |
| High school or less | 482 | 30.7 | 30.6 | 25.4-35.7 | 0.03 | 1.06 | 0.87-1.30 | 0.55 | 43.0 | 37.4-48.6 | <0.0001 |
| Some college | 546 | 31.4 | 37.8 | 33.6-42.1 |  | 1.52 | 1.13-2.05 | 0.01 | 46.2 | 41.8-50.5 |  |
| ≥Completed college | 1015 | 36.5 | 38.0 | 34.8-41.3 |  | 1.00 | Ref. | -- | 60.5 | 57.2-63.7 |  |
| Missing<sup>s</sup> | 31 | 1.4 |  |  |  |  |  |  |  |  |  |
| **Household** |  |  |  |  |  |  |  |  |  |  |  |
| **Number of children ≤12 years** |  |  |  |  |  |  |  |  |  |  |  |
| 1 | 1059 | 50.8 | 36.8 | 33.5-40.1 | 0.08 | 1.00 | Ref. | -- | 51.8 | 48.4-55.3 | 0.01 |
| 2 | 751 | 34.7 | 37.3 | 33.2-41.3 |  | 0.99 | 0.86-1.14 | 0.87 | 53.1 | 48.9-57.4 |  |
| 3 or more | 264 | 14.4 | 29.5 | 0.0-1.8 |  | 0.86 | 0.68-1.08 | 0.20 | 40.3 | 32.9-47.7 |  |
| **Income (USD)** |  |  |  |  |  |  |  |  |  |  |  |
| <\$25,000 | 331 | 20.3 | 29.7 | 23.9-35.5 | <0.0001 | 0.82 | 0.67-1.00 | 0.05 | 43.9 | 37.4-50.4 | <0.0001 |
| \$25,000-\$49,999 | 472 | 24.6 | 31.8 | 26.9-36.7 | | 0.84 | 0.72-0.98 | 0.03 | 41.6 | 36.5-46.7 | |
| \$50,000-\$99,999 | 587 | 27.4 | 34.5 | 30.2-38.8 | | 0.84 | 0.66-1.07 | 0.15 | 52.3 | 47.7-56.9 | |
| ≥\$100,000 | 617 | 23.9 | 48.6 | 44.1-53.2 | | 1.00 | Ref. | -- | 68.3 | 64.2-72.4 | |
| Missing<sup>s</sup> | 67 | 3.8 |  |  |  |  |  |  |  |  |  |
| **Region** |  |  |  |  |  |  |  |  |  |  |  |
| Northeast | 550 | 15.7 | 42.1 | 37.4-46.8 | 0.02 | 1.00 | Ref. | -- | 52.5 | 47.7-57.2 | 0.29 |
| South | 684 | 39.0 | 34.3 | 30.2-38.3 |  | 0.87 | 0.74-1.02 | 0.08 | 48.3 | 44.0-52.7 |  |
| Midwest | 442 | 21.0 | 30.3 | 25.7-34.9 |  | 0.76 | 0.63-0.91 | <0.01 | 47.6 | 42.5-52.7 |  |
| West | 398 | 24.3 | 39.4 | 33.9-44.9 |  | 0.96 | 0.81-1.13 | 0.65 | 55.7 | 50.0-61.4 |  |

Abbreviations: CI = confidence interval; IQR = interquartile range; USD = US dollars.
\*Survey weights applied to sample to represent US population of parents by race, ethnicity, sex, education and region.
^Weighted percentages are prevalence estimates of US parents reporting vaccination plans for their youngest child.
†p-values from Rao adjusted Pearson Chi-squared tests to compare expected to observed frequencies among groups by characteristic for parent's willingness to vaccinate their youngest child (i.e. whether willing to vaccinate youngest child differed by sex of the child, etc.);
§Categories are not presented in the table as they yielded unreliable standard error estimates.
\*\*Child's race/ethnicity excluded from adjusted models due to collinearity with parent's race/ethnicity.
^^Parents identifying as transgender were grouped with their identified gender.

Overall, 50.6% of parents said they would allow their youngest child to be tested for COVID-19 at school/daycare if required, 33.5% said they would not allow school-based testing and 14.4% said they didn’t know (1.5% did not answer). In univariable analyses, parents of children <2 years of age, parent of children without health insurance and parents of children not attending in-person school were less willing to allow school-or daycare-based COVID-19 testing (Table 1). We also observed that non-Hispanic Black parents, those who were 18-24 years of age, parents who had more than 2 children in the home, those who had not completed college and parents with income <$50,000 were less likely to report that they would allow their child to be tested for COVID-19 in school or daycare settings (Table 1).

## DISCUSSION

COVID-19 testing among U.S. children has been low to date, in our study, only 36% of US parents reported their child had ever been tested. Female and Asian parents were least likely to report their child having been tested for COVID-19, while parents of children with health insurance and who were attending school were more likely to report testing. We also found the most preferred COVID-19 testing location was the child’s pediatrician’s office. Finally, only half of parents in our survey reported they would allow their child to be tested at school or daycare which also varied by demographic characteristics.

To maximize uptake of COVID-19 testing among unvaccinated children, an important testing implementation strategy may involve the ability for providers, laboratories, and parents to be able to authorize and easily provide and disclose certified proof of test results for children (e.g., via a national app) to entities (e.g., schools), thus allowing flexibility and the ability for parents to choose a preferred testing provider and venue that meets basic standards for COVID-19 testing. In addition, free pediatric COVID-19 testing across the US and efforts to increase parental awareness of no-fee testing venues are also needed to ensure greater access and wider uptake among children without health insurance and others facing testing barriers.

There are several limitations to our analysis. Our survey focused on children ≤12 years of age in order to collect information about younger children and does not provide information about adolescents. Survey data were self-reported and subject to recall, response and social desirability bias. The survey was weighted to reflect the US population of parents based on 2019 census estimates. However, it was conducted online, and therefore, excluded parents without access to the internet.

Our study provides important information about children who have been tested for COVID-19 thus far in the epidemic and those who may be missed for testing. It also includes novel information about venues where parents prefer to seek testing for their children. Given that a COVID-19 vaccine for children may not be approved for pediatric population until late in 2021(9), and that COVID-19 vaccine hesitancy among parents may be significant(10), testing will remain critical for identifying children with infection and as a public health intervention to contain infections. As such, our data can inform strategies to increase testing coverage and acceptability among US children.

## Data Availability

Survey data will be available upon request.

## References

1. Amercian Academy of Pediatrics (AAP) and Children’s Hospital Association. Children and COVID-19: State Data Report Version: May 13, 2021. https://services.aap.org/en/pages/2019-novel-coronavirus-covid-19-infections/children-and-covid-19-state-level-data-report/. Accessed: May 10, 2021.

2. Goodman B. Cases in children helping the surge of COVID variants. WebMD Health News. Accessed April 9, 2021. https://www.webmd.com/lung/news/20210402/cases-in-children-helping-surge-of-covid-variants.

3. Felsenstein S, Hedrich CM. SARS-CoV-2 infections in children and young people. Clin Immunol. 2020;220:108588.

4. Buonsenso D, Munblit D, De Rose C, Sinatti D, Ricchiuto A, Carfi A, et al. Preliminary evidence on long COVID in children. Acta paediatrica. 2021.

5. Leidman E, Duca LM, Omura JD, Proia K, Stephens JW, Sauber-Schatz EK. COVID-19 Trends Among Persons Aged 0-24 Years - United States, March 1-December 12, 2020. MMWR Morb Mortal Wkly Rep. 2021;70(3):88–94.

6. Sisk B, Cull W, Harris JM, Rothenburger A, Olson L. National Trends of Cases of COVID-19 in Children Based on US State Health Department Data. Pediatrics. 2020;146(6).

7. Standard defintions - American Association for Public Opinion Research (AAPOR). https://www.aapor.org/AAPOR_Main/media/publications/Standard-Definitions20169theditionfinal.pdf. Accessed: January 5, 2021.

8. US Census Bureau (USC). Census QuickFacts (2019). https://www.census.gov/quickfacts/fact/table/US/PST045219. Accessed January 5, 2021.

9. Centers for Disease Control and Preventio (CDC). Summary Document for Interim Clinical Considerations for use of COVID-19 Vaccines Currently Authorized in the United States. Atlanta, GA: CDC; 2021. https://www.cdc.gov/vaccines/covid-19/info-by-product/clinical-considerations.html. Accessed: May 10, 2021.

10. Teasdale CA, Borrell LN, Kimball S, Rinke ML, Rane M, Fleary SA, et al. Plans to vaccinate children for COVID-19: a survey of US parents. medRxiv pre-print 2021.

